# Treatment outcomes of tuberculosis among people living with HIV/AIDS: A comparative assessment of primary and tertiary healthcare centres in Nasarawa State, Nigeria

**DOI:** 10.1101/2025.02.25.25322853

**Authors:** Benjamin Mari Aya, Ismail Ndalami Salihu, Dalhatu Muhammad Ahmad, Dinfa Dombin Tyem, Ovye Stephen Mari, Namo David Alu, Benjamin Nasara Joseph, Faisal Shuaib

**Author notes:** Corresponding author (BMA).

## Abstract

One of the Sustainable Development Goals for 2030 is to end the global TB epidemic. Translating this laudable target to reality requires strengthening collaborative tuberculosis/human Immunodeficiency Virus (TB/HIV) care at all levels to end TB in Nigeria. This study assessed the treatment outcomes of TB among people living with HIV (PLWH) and compared the treatment outcomes of TB between primary and tertiary healthcare facilities in Nasarawa State, Nigeria.

This facility-based retrospective survey of TB patients living with HIV was conducted in two tertiary healthcare facilities and a primary healthcare centre (PHC). Records of eligible patients who completed treatment between January 2016 and December 2019 were abstracted using a data pro forma. Treatment outcomes were categorized as either successful or unsuccessful. Descriptive and inferential analyses were conducted, in addition to multivariate logistic regression.

A total of 959 patients with a mean age of 33±14 years were evaluated; there were 847 adults (88.3%), and the proportion of females was 499 (52%). The proportion of patients who completed TB treatment was 26%; 20.5% were cured, while the mortality rate was 9.2%. The treatment success rate (TSR) was 46.5%; tertiary healthcare facilities had the highest unsuccessful treatment rates compared to the PHC. The TSR declined steadily from 68.8% in 2016 to 57.6% in 2019. Being treated at a tertiary healthcare facility (AOR=3.6, CI; 2.0-6.6) predicted successful treatment outcomes of TB among PLWH.

TB’s overall TSR among PLWH was low. Within the outcome categories, the proportion of patients who completed treatment and those cured compared better with unfavourable outcomes. The rate of unsuccessful TB treatment is highest in tertiary healthcare facilities compared to PHC. However, being treated in the tertiary healthcare facility predicted successful treatment outcomes, while treatment failure, death, and loss to follow-up were highest in the tertiary healthcare facilities.

## Background

The age-long battle to end the scourge of tuberculosis (TB) has been frustrated by the emergence of the human Immunodeficiency virus (HIV) [1]. HIV/TB-coinfection constitutes an unresolved public health challenge in sub-Saharan Africa and the entire world [2]. Nigeria is the 4th out of 30 countries with a high TB burden, with an incidence rate of 219 per 100,000 population. The estimated incidence of TB among people living with HIV (PLWH) is 34 per 100, 000 and the mortality is 21 per 100,000 [3]. The risk of developing active TB among PLWH falls between 16-27 times more than those without HIV, thereby amplifying the spread of TB [1, 4]. This suggests that the TB elimination target set for 2050 could be compromised if the dual burden of TB/HIV coinfection is not controlled [5].

In 2015 alone, the proportion of patients with TB and HIV coinfection in Nasarawa State was 35.8%, far above the Nigeria average of 12% [6, 7]. Additionally, an estimated 40% of TB clients who returned for retreatment in the same state were coinfected with HIV [8]. This is an issue of great concern since Nasarawa State has a high HIV prevalence of 2.0% according to the recent Joint United Nations Program on HIV/AIDS (UNAIDS) country estimates [9]. Although PLWH could have other diseases, TB is by far the leading single cause of death requiring a deliberate and concerted effort to halt these dual epidemics beyond the broader strengthening of HIV service delivery [3].

Furthermore, evidence suggests that the low treatment success rate of TB is attributed to weaknesses in patient management, which results in poor patient adherence and approximately a 30% loss to follow-up [10, 11]. This is particularly worrisome since concomitant treatment of TB and HIV is faced with the challenges of higher pill burden and overlapping drug toxicities, which may directly affect adherence to the drug regimen and, ultimately, treatment outcomes of TB. Fan and colleagues pointed out that this problem is underpinned by limited resources and is worsened in developing countries such as Nigeria, where already strained health service delivery systems continue to impede current control measures against TB [12, 13].

The goal of the TB program in Nigeria is to reduce TB/HIV-associated morbidity and mortality through collaboration between the National Tuberculosis and Leprosy Control Program (NTBLCP) and the National Agency for the Control of AIDS (NACA) [14]. To achieve the goals of this strategy amidst the poorly equipped laboratory facilities at all levels of the healthcare delivery system requires the decentralization of TB and HIV care centres, adoption of the primary healthcare model for TB/HIV care, integration of TB/HIV care and mobilization of highly skilled professionals to improve capacities of health workers through intensifying the task-shifting, task-sharing structures, and monitoring that would otherwise improve treatment outcomes [15]. Nasarawa State, like other states in northern Nigeria, relies on task shifting for both TB and HIV care. In primary healthcare centres (PHCs), however, there is underperforming access to diagnosis, lack of involvement and coordination between managers, vertical management, and high turnover of professionals – which arguably contributes to the poor performance of TB control in PHC [16]. Although completion of treatment is monitored primarily by the Public Health Unit, information on treatment outcomes of TB is sparingly reported [17]. Worse still, poor treatment outcomes have serious consequences, including infectivity and the development of drug-resistant Mycobacterium TB [18].

While evidence has demonstrated treatment outcomes of TB at the facility level, no single study has compared the performance in terms of treatment outcomes of TB among PLWH between primary and tertiary healthcare facilities. This study sought to answer what are the treatment outcomes of TB among PLWH in Nasarawa State, Nigeria? How do TB treatment outcomes differ between primary and tertiary healthcare facilities? What are the predictors of successful treatment outcomes among TB-treated PLWH in Nasarawa State, Nigeria?

## Methods

### Study Setting

The study sites were TB treatment centres of Federal Medical Centre (FMC), Keffi; Dalhatu Araf Specialist Hospital (DASH), Lafia; and Evangelical Reformed Church of Christ Hospital (ERCC) Hospital Alushi, Nassarawa Eggon. The FMC Keffi and DASH Lafia are tertiary healthcare facilities, while the ERCC Hospital is a faith-based PHC. These facilities account for over 50% of all TB cases in Nasarawa State. DASH is situated in the state capital, Lafia, in the southern part of the state. In contrast, ERCC Hospital Alushi, Nassarawa Eggon is situated in the northern part of the state. FMC Keffi, on the other hand, is located in the western part of the state. These hospitals are known for their long-term history of TB treatment with support from donor agencies. They offer integrated TB/HIV care and support programs based on the exemption policy on TB and antiretrovirals [8]. Moreover, they provide directly observed treatment short-course (DOTS) services and are reference centres for diagnosing and treating TB.

### Study Population

All patients diagnosed with TB, irrespective of the diagnostic criteria of the infection with background HIV disease at initiation of therapy, and who had received and completed a course of treatment.

### Study Design

This was a cross-sectional facility-based retrospective study of TB patients records from January 2016 to December 2019 in three DOT TB treatment facilities in Nasarawa State, Northcentral Nigeria to examine TB treatment outcomes.

### Inclusion & Exclusion Criteria

Males and females aged 1 month and above who were registered in healthcare facilities for pulmonary or extrapulmonary TB were included in the study. The research only includes clients who completed their treatment between January 2016 and December 2019. Pregnant women and those with multidrug-resistant TB were excluded from the research.

### Study Instrument & Data Collection

The research employed a data proforma to elicit information relevant to the study. The instrument is divided into three sections. The first section elicited the demographic characteristics of the patients, while the second section centred on the clinical characteristics of the patient. The third section investigated the treatment outcomes of the patients. Data were extracted manually from healthcare facility TB registers and treatment cards of patients who accessed TB treatment and completed therapy between January 2016 and December 2019 using a structured checklist before transferring to the Microsoft Excel spreadsheet. The procedure followed is presented in Fig 1.

**Fig 1:**
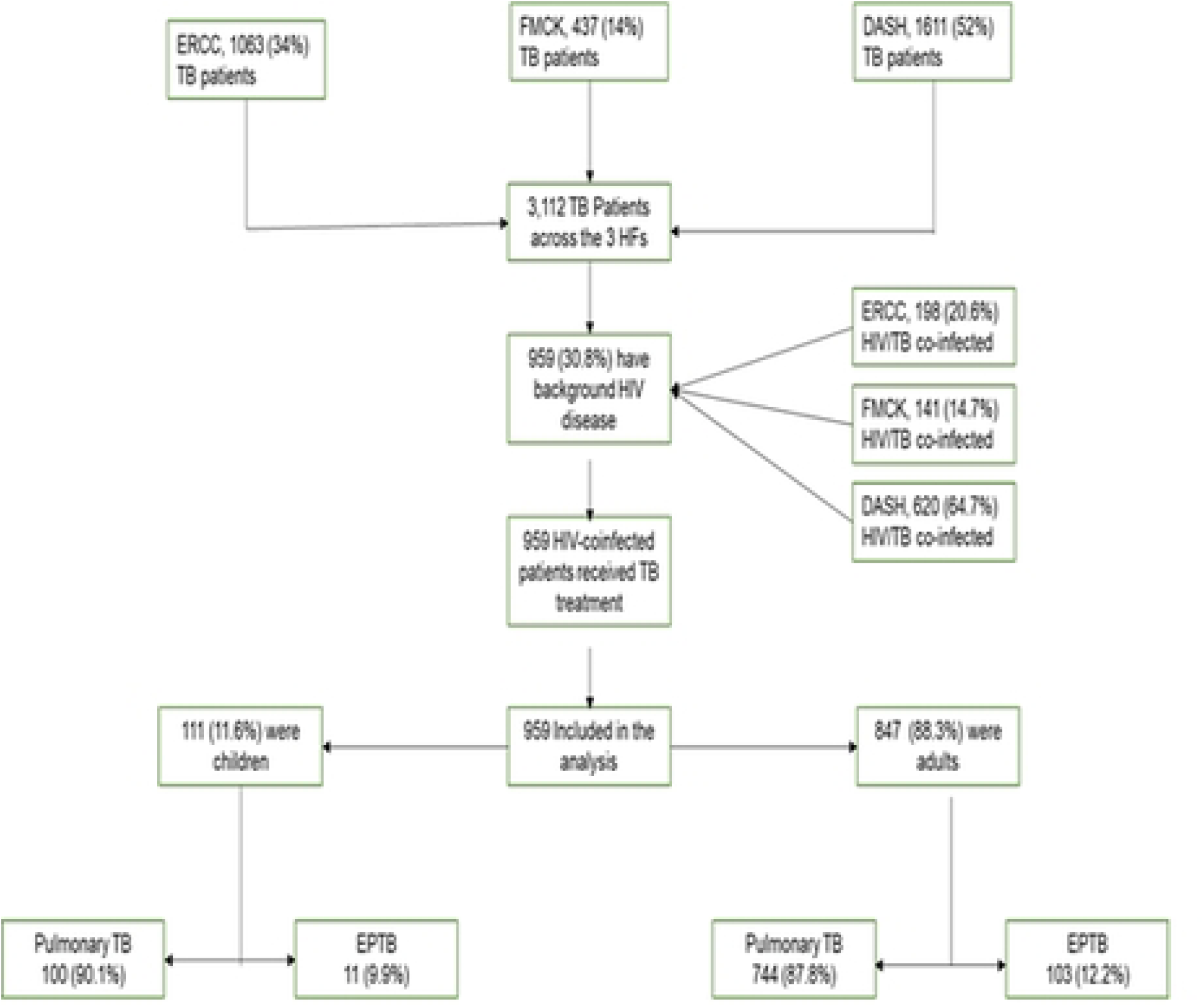
Data extraction flow diagram

### Outcome Measures

Treatment outcome is defined as cured, treatment completed, default, death, relapse, retreatment, not evaluated, and treatment failure. In this study, successful treatment was conceptualized as those who were cured or completed their treatment. Other outcomes, such as default, death, or treatment failure were defined as unsuccessful treatment.

### Statistical Analysis

Standard exploratory analysis was used to describe the demographic and clinical characteristics of the patients. Continuous data were tested for normality using the Shapiro-Wilk test. Means with standard deviation (±SD) were reported for continuous variables that showed a normal distribution, while medians with interquartile ranges were presented for continuous variables that violated the assumption of normality. Bivariate analysis was used to determine factors associated with treatment outcomes. For comparison between groups, Pearson’s chi-square test of independence (*X*^2^) and Fisher’s exact test (FET) were performed for variables at the nominal level of measurement. Furthermore, clinically relevant variables that plausibly influenced treatment outcomes were fitted in a multivariate logistic regression model to identify independent predictors of treatment outcomes. Comparisons were made across the different healthcare facilities to determine their similarities and differences. The groups found to be similar were combined for “state-level treatment outcomes” before merging the two tertiary healthcare facilities for comparison with the PHC, ERCC Hospital. Statistical tests were two-sided, with alpha defined at p<0.05. All statistical analyses were performed using the Statistical Package for the Social Science (SPSS) version 20 for Windows.

### Ethical Statement

The conduct of this research was approved by the Health Research and Ethics Committee of the Federal Medical Centre Keffi with the reference number FMC/KF/HREC/384/20.

### Confidentiality

Anonymity was maintained during data collection to ensure that patient information was treated with the utmost confidentiality.

## Results

A total of 3,112 records were abstracted across all the selected healthcare facilities, within the study period, out of which 959 were included in the final analysis. The mean age was 33±14 years, and adults (847, 88.3%) constituted the highest proportion of the TB patients evaluated. Females were the majority in the study (499, 52%) (Table 1).

**Table 1:**
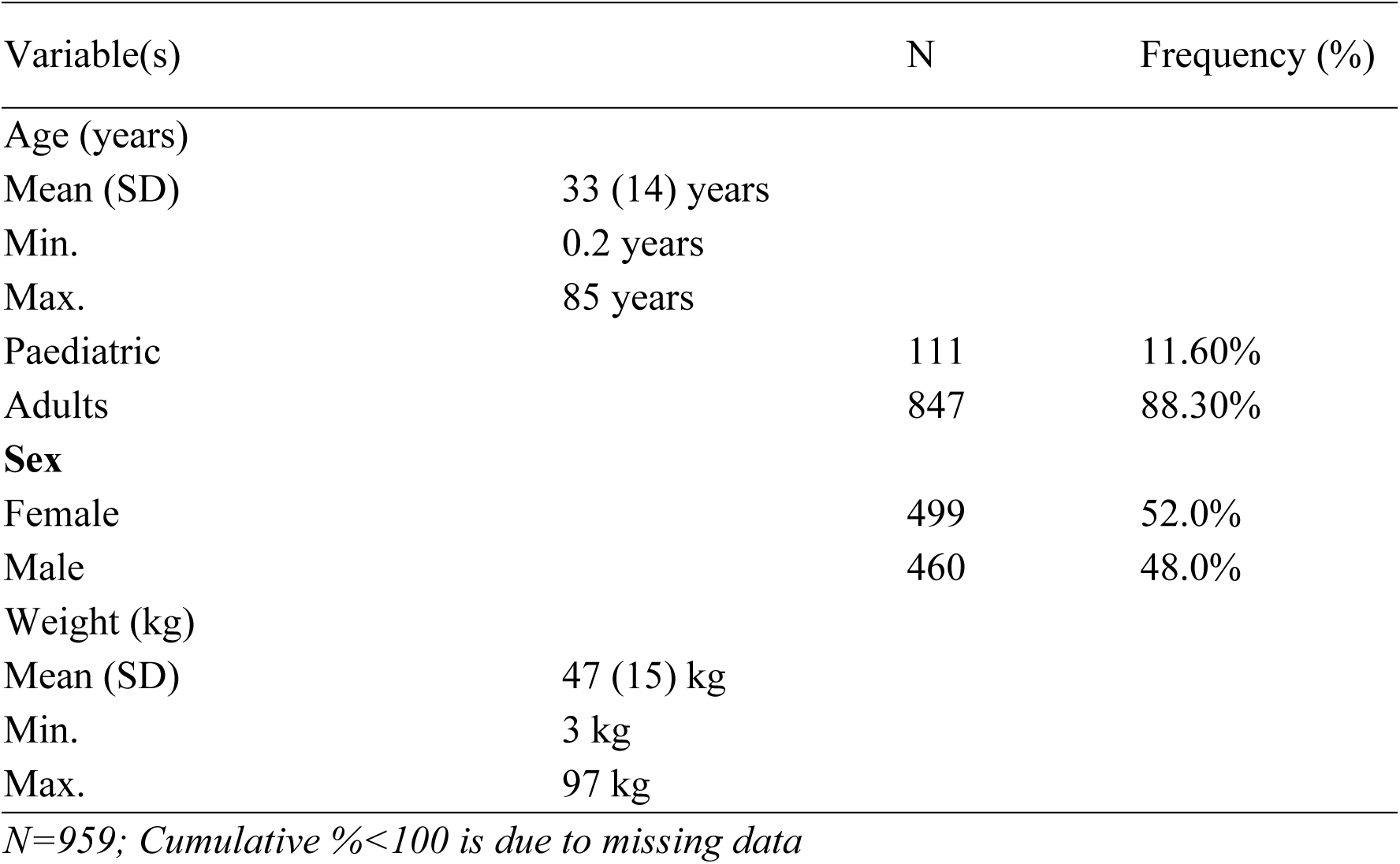
Demographic characteristics of patients.

More females (399, 80%) and paediatrics (99, 89%) (p<0.05) were treated in tertiary healthcare facilities. Most of the patients treated in the tertiary healthcare facility had previous knowledge of their HIV status [747 (79%), p >0.05] with an estimated [738 (82%), p < 0.001] active on antiretroviral therapy. Diagnostic criteria in the PHC relied on Acid Fast Bacilli [60 (100%), p <0.001] and clinical judgment of the clinician [29 (100%), p <0.001], whereas chest X-ray [198 (87%), p <0.001] and GeneXpert [192 (72%), p <0.001] were predominantly used for diagnosis in tertiary healthcare facilities (Table 2).

**Table 2:** Demographic and clinical characteristics based on facility type.

| Variable(s) | Categories | Facility type |  | p-value |
| --- | --- | --- | --- | --- |
|  |  | Primary | Tertiary |  |
| Sex | Female | 100(20%) | 399(80%) | 0.629 |
|  | Male | 98(21%) | 362(79%) |  |
| | $X^2=0.234$; $df=1$ | | | |
| Age | Paediatric | 12(11%) | 99(89%) | 0.006* |
|  | Adults | 186(22%) | 661(78%) |  |
| | $X^2=7.44$; $df=1$ | | | |
| Previously known HIV Status | No | 2(13%) | 14(88%) | 0.546 |
|  | Yes | 196(21%) | 747(79%) |  |
| | $FET=0.659$; $df=1$ | | | |
| Active on ART | No | 40(64%) | 23(37%) | <0.001* |
|  | Yes | 158(18%) | 738(82%) |  |
| | $X^2=75.6$; $df=1$ | | | |
| Diagnostic criteria | Acid Fast Bacilli | 60(100%) | 0(0%) | <0.001* |
|  | Chest X-ray | 31(13%) | 198(87%) |  |
|  | GeneXpert | 76(28%) | 192(72%) |  |
|  | Clinical | 29(100%) | 0(0%) |  |
| | $X^2=220$; $df=3$ | | | |
| Anatomical site of TB | Pulmonary | 190(23%) | 654(77%) | <0.001* |
|  | Extrapulmonary | 8(7%) | 106(93%) |  |
| | $X^2=14.7$; $df=1$ | | | |
| $X^2=Chi\ Square\ test$; $FET=Fisher's\ Exact\ Test$; $df=degree\ of\ freedom$; *statistical sig. ($p<0.05$) | | | | |

Overall, slightly above one-quarter of the patients evaluated completed treatment (26%); 20.5% were cured, while the mortality rate was 9.2%. As many as 14.6% of the clients defaulted/lost to follow-up, whereas 3% were not evaluated (Fig 2).

**Fig 2:**
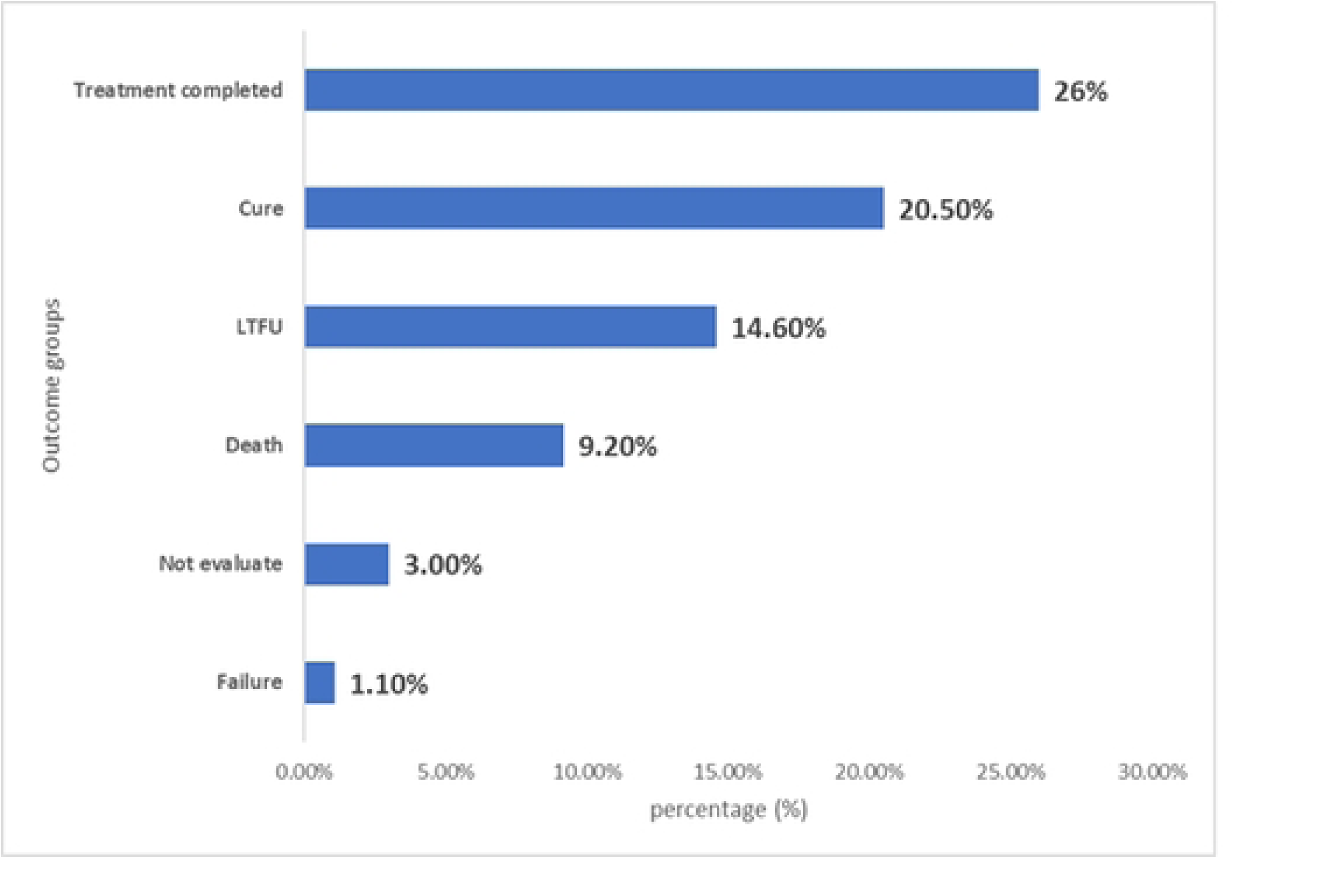
Treatment outcomes of TB among PLWH

The overall state-level treatment success rate was 46.5% (Fig 3).

**Fig 3:**
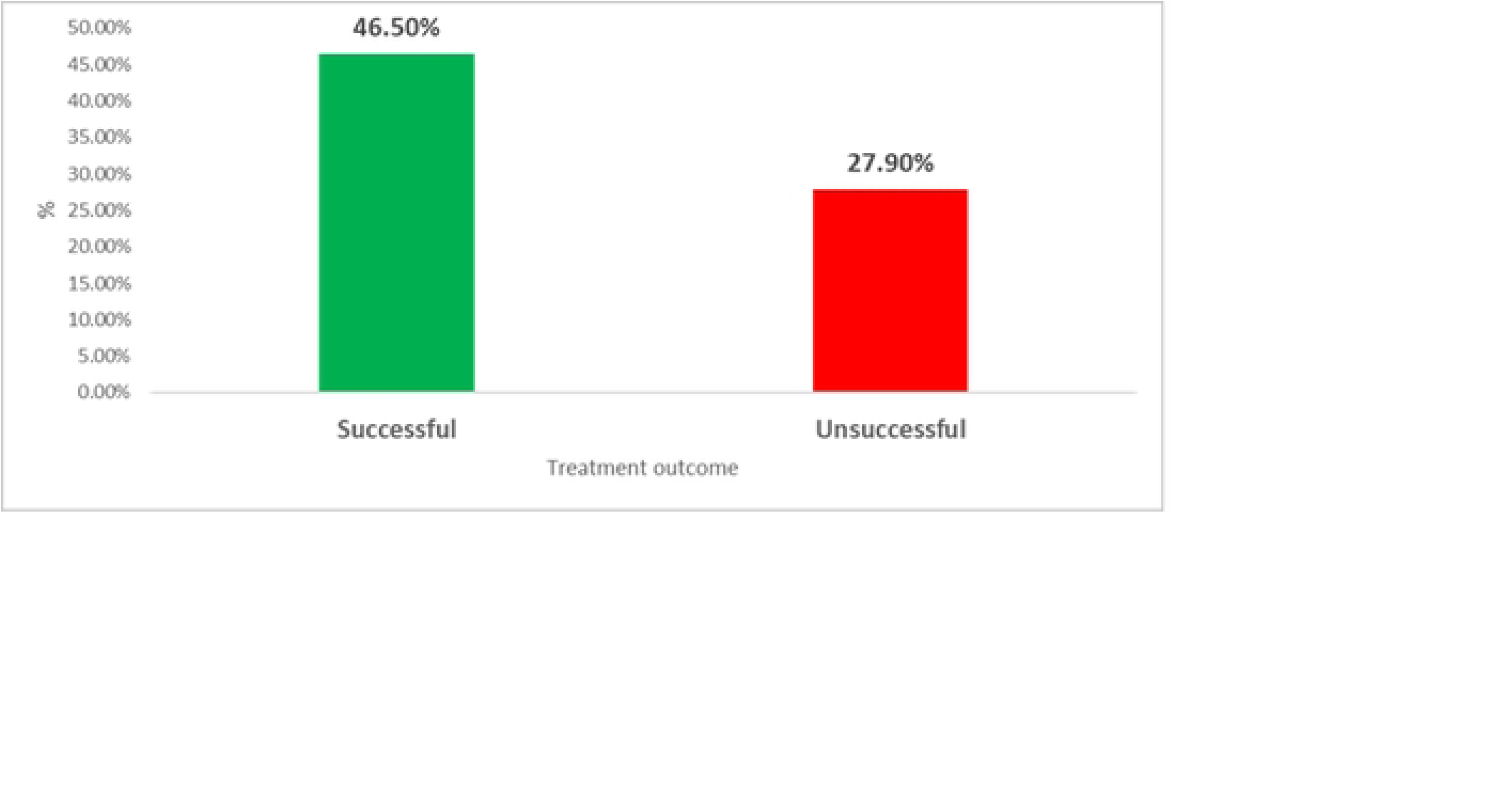
Treatment success rate of TB among PLWH

The trend of TB treatment outcomes illustrates a steady decline in the treatment success rate over time, with a peak of 68.8%, which dropped to a lower rate of 57.6% in 2019. This is in light of a steady increase in the unsuccessful rate over time, with a peak of 42.4% in 2019 (Fig 4).

**Fig 4:**
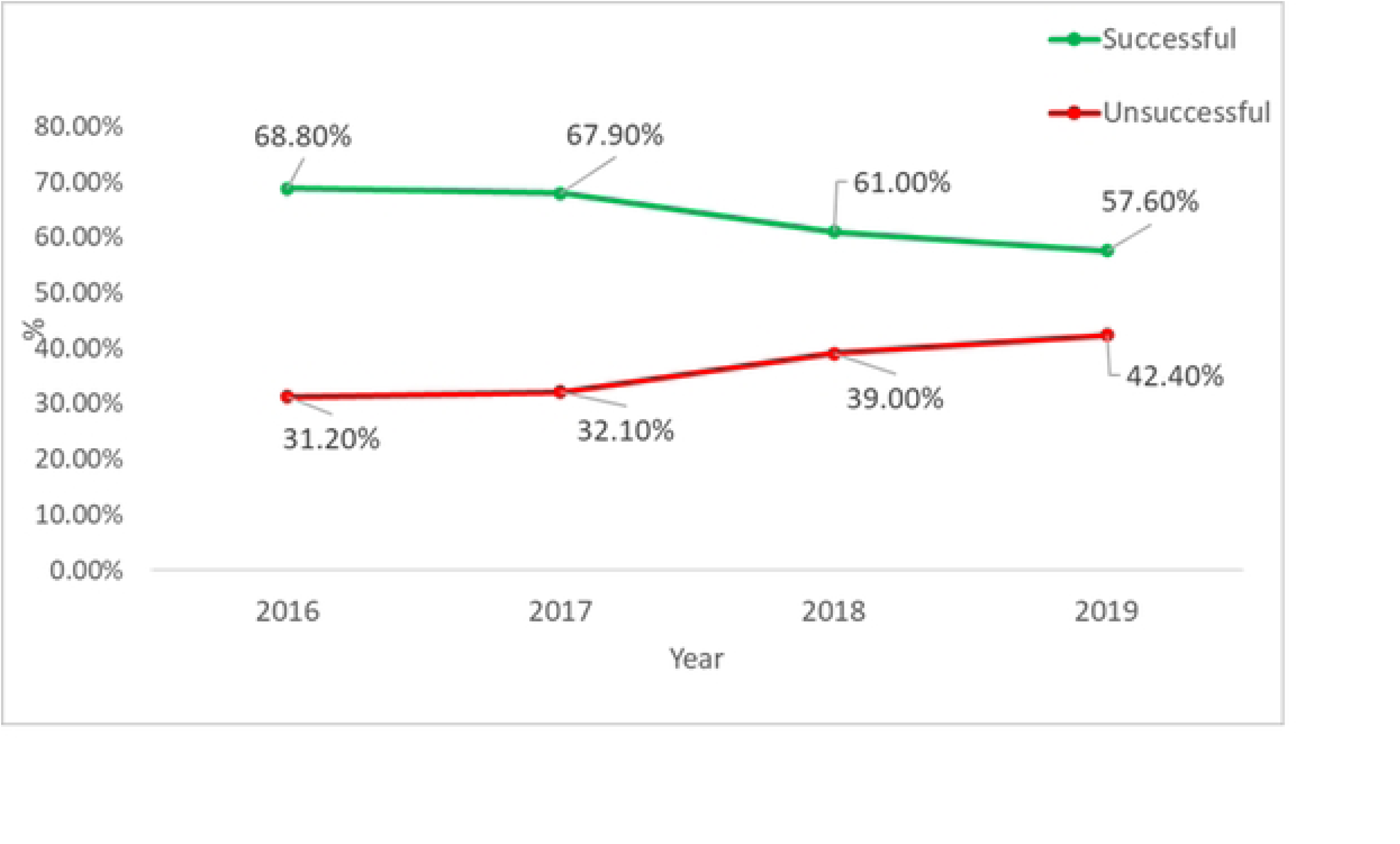
Trends of TB treatment outcomes among PLWH

Outcome groups of TB treatment statistically differ significantly [FET (df) = 151.6(5), p < 0.001] across the primary and tertiary healthcare facilities. The cure rate (59.9% versus 40.1%) was highest in the PHC. However, rates for treatment failure (63.6% versus 36.4%), death (97.7% versus 2.3%), loss to follow-up (82.9% versus 17.1%), and patients not evaluated (86.2% versus 13.8%) were observed in the tertiary healthcare facility (Fig 5).

**Fig 5:**
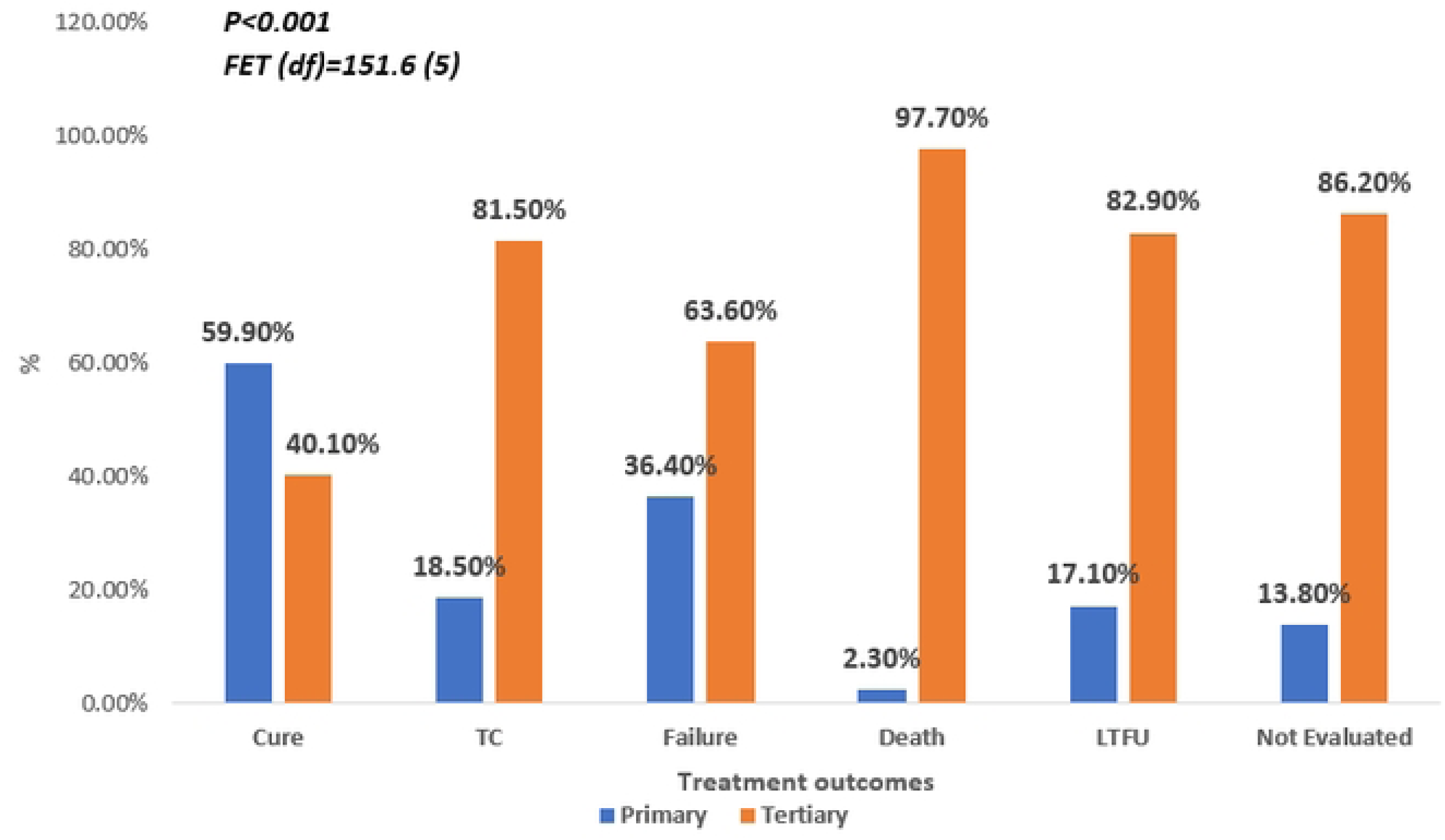
Treatment outcomes of TB based on facility type

An overview of the treatment outcomes of TB among PLWH demonstrated that PHC (ERCC Hospital, Alushi) had a higher successful treatment rate and unsuccessful outcomes were greater for tertiary healthcare facilities (DASH, Lafia and FMC, Keffi) (Fig 6).

**Fig 6:**
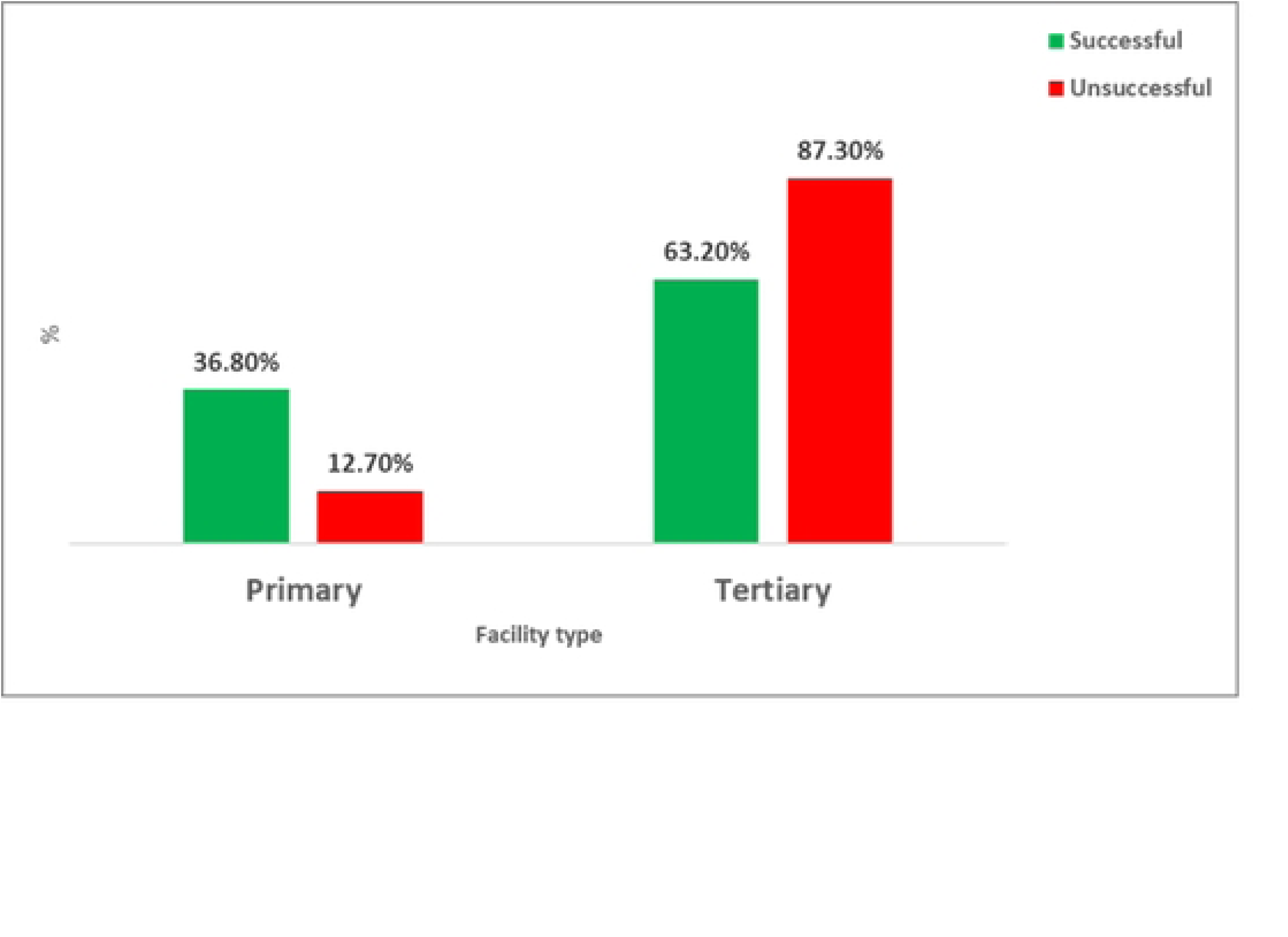
Treatment success rate of TB among PLWH

When the TB treatment outcomes among PLWH were compared with demographic and clinical characteristics, there was no significant difference with sex and previous knowledge of HIV status (p ≥0.05). There was a statistically significant difference with age (p = 0.025), being active on antiretroviral therapy (p < 0.001), diagnostic criteria (p < 0.001), and anatomical site of TB infection (p = 0.012). Specifically, females (28%) were more likely to have a cure outcome for TB than males, similar to patients with prior knowledge of their HIV status (28%), which was statistically insignificant (p>0.05) (Table 3).

**Table 3:**
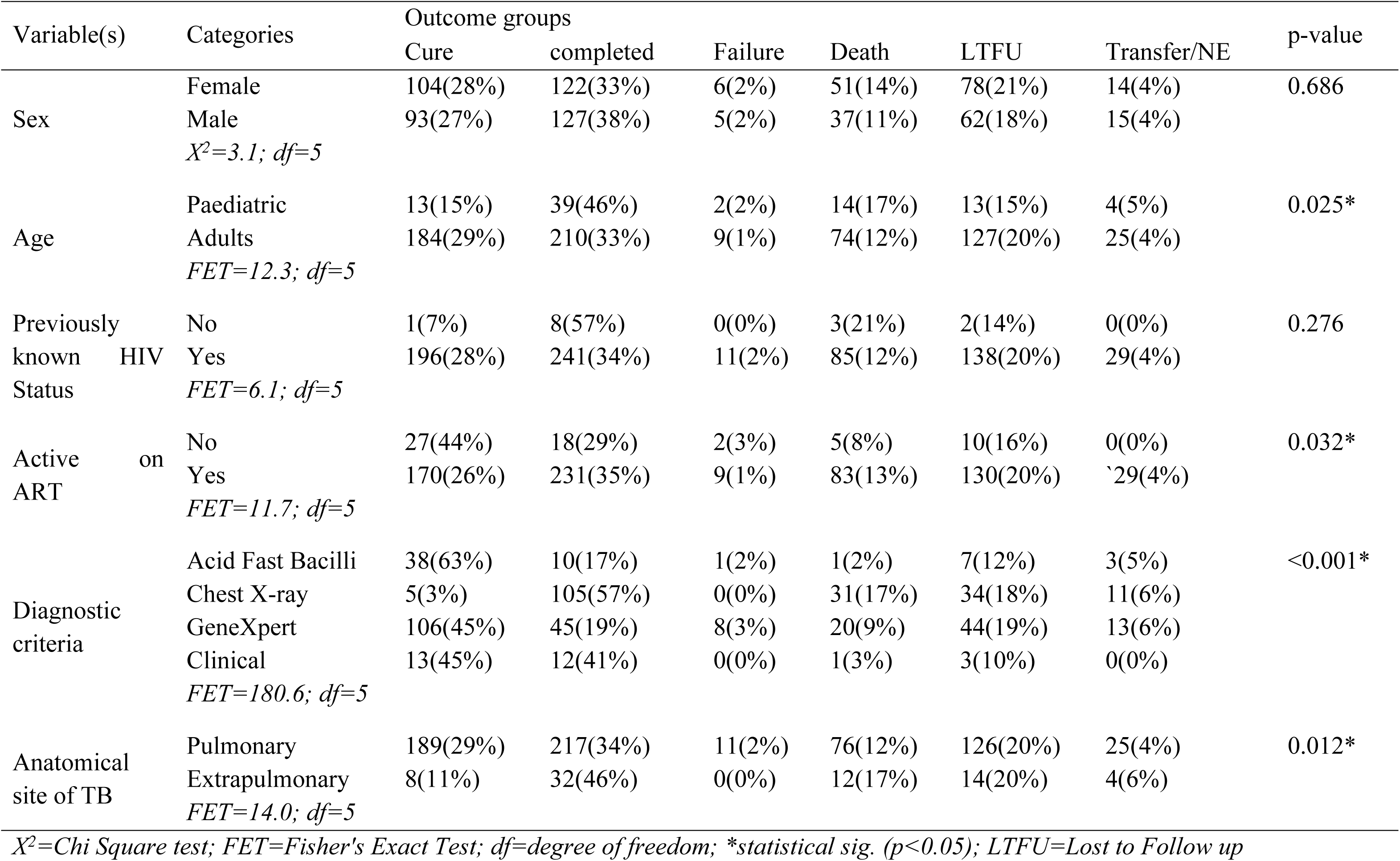
Factors associated with treatment outcomes of TB (N=959)

Table 4 below illustrates the interaction between outcome categories and facility type. The PHC had the highest cure rate of 724 (71.5%), the lowest mortality rate of 4 (2.9%), loss to follow-up of 105 (25.9%), and the proportion of patients not evaluated of 21 (19.4%). This interaction was statistically significant (p <0.001).

**Table 4.** Comparison of treatment outcome groups based on facility type.

| Variable(s) | Facility type |  | X<sup>2</sup> (df) | P-value |
| --- | --- | --- | --- | --- |
|  | Primary | Tertiary |  |  |
|  | N (%) | N (%) |  |  |
| Outcome categories |  |  |  |  |
| Cure | 724(71.5%) | 288(28.5%) | 606.1 (5) | <0.001* |
| Treatment completed | 187(24.7%) | 569(75.3%) |  |  |
| Treatment failure | 19(55.9%) | 15(44.1%) |  |  |
| Death | 4(2.9%) | 136(97.1%) |  |  |
| Loss to follow-up | 105(25.9%) | 301(74.1%) |  |  |
| Not evaluated | 21(19.4%) | 87(80.6%) |  |  |

*X<sup>2</sup>=Chi Square test; df=degree of freedom; \*statistical sig. ( $p < 0.05$ )*

The chi-square test of Independence revealed evidence for the relationship between diagnostic criteria and treatment outcome of TB among PLWH (p = 0.002). More males [220 (65%), p = 0.202] had successful treatment outcomes than females. Likewise, adults [394 (63%), p = 0.794] were more likely to have a successful treatment outcome than paediatrics. Patients who had no prior knowledge of HIV status [9 (64%), p = 0.887], as well as those not active on antiretroviral therapy [45 (73%)], had the highest treatment success rate compared to their counterparts (Table 5).

**Table 5:**
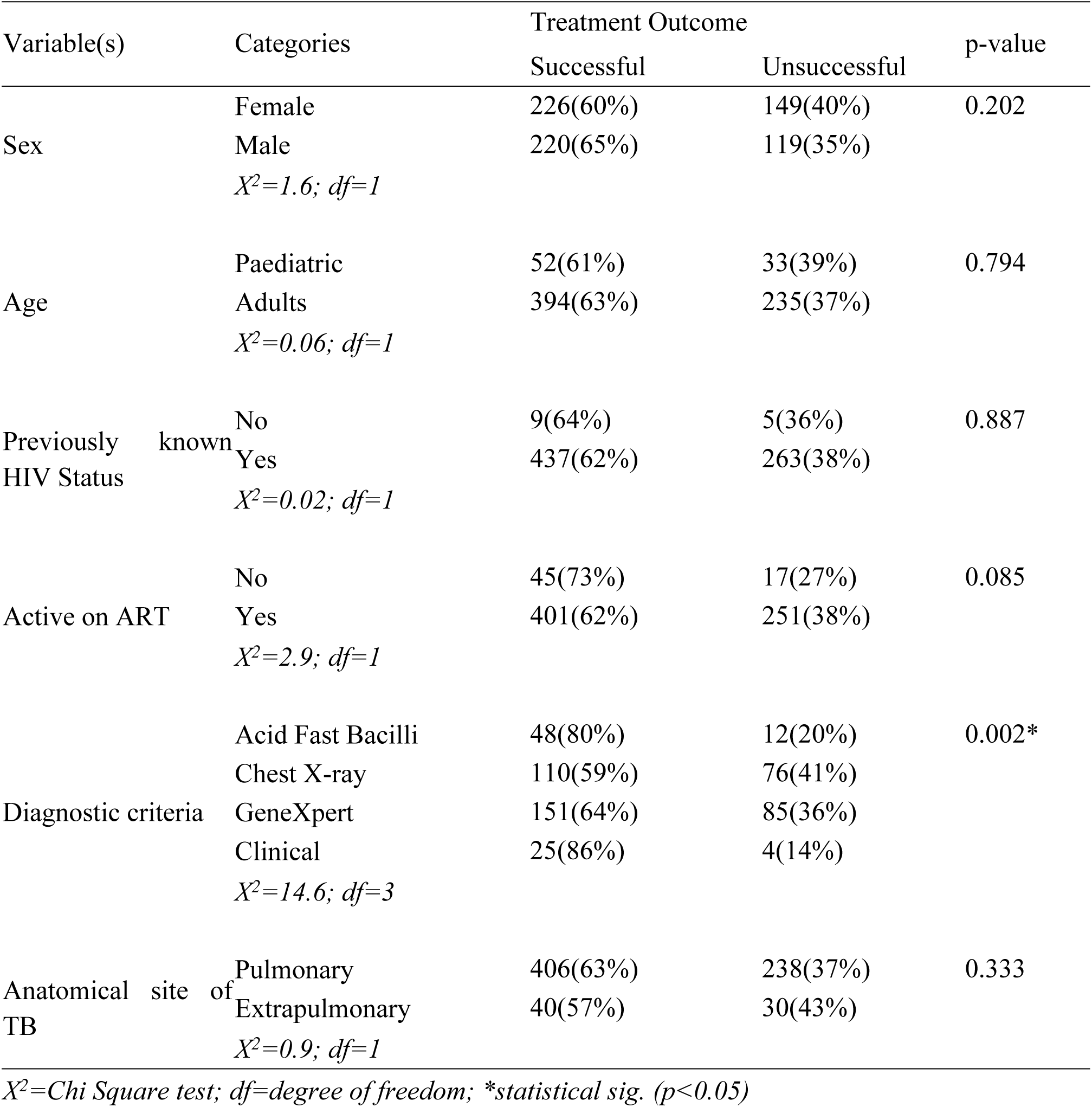
Demographic and clinical characteristics based on treatment outcomes (N=959)

A binary logistic regression model was fitted to identify independent predictors of treatment success. After adjusting for sex, age, active on antiretroviral therapy, anatomical site of TB infection, and previous TB treatment, in a multivariate regression model, being treated in a tertiary healthcare facility (aOR [3.6 (2.0-6.6)], p<0.001) predicted successful treatment outcomes of TB among PLWH (Table 6).

**Table 6:**
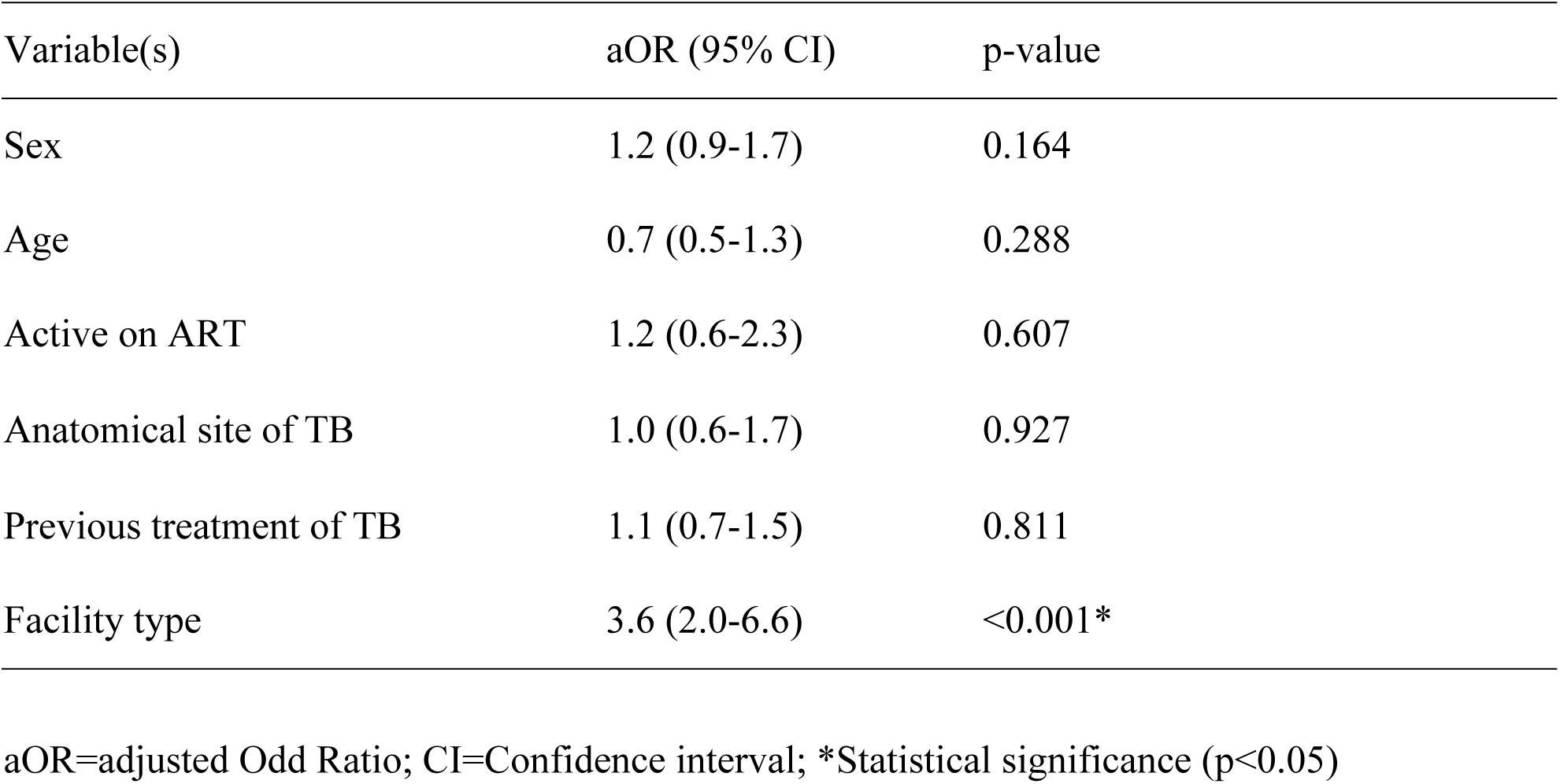
Predictors of treatment outcomes.

## Discussion

This study demonstrated an overall treatment success rate (TSR) of 46.5%, slightly less than the 57% reported in Kuala Lumpur [19]. Furthermore, the four-year mean TSR in this study is lower than the reports obtained from other regions of Nigeria: North (83.7%), Southeast (75.7% to 86%), and Southwest (83.5%) [20, 21, 22, 23]. Findings from a systematic review and meta-analysis across six European countries revealed a national pooled estimate of 68.1%, which was also higher than the TSR found in this study [24]. Even though treatment outcomes of TB among PLWH have increased significantly over recent decades with the advent of antiretroviral therapy (ART), the lower TSR in this research cannot be unconnected with the fact that PLWH were the cardinal focus of this research. A previous study showed that treatment success rates in hospitals in Nigeria varied widely between 43.7% and 86% [25]. The success rate for the treatment of TB among PLWH has increased significantly over the past several decades due to advances in ART. According to a review published in 2018, the overall TSR for TB among PLWH is 75%, and it may be as high as 85% in some settings where there is good access to ART and TB care [1]. It is worth noting that the TSR varies depending on several factors, including the severity of TB infection, the presence of drug-resistant TB, and the patient’s overall health. Studies from Ethiopia and Ghana showed a higher TSR for TB among PLWH compared to what is observed in this study [26, 27].

The PHC achieved a statistically significant outcome within the cure category [118 (59.9%), p <0.001] compared to the tertiary healthcare facilities. Conversely, the tertiary healthcare facilities achieved a better outcome for treatment completed [203 (81.5%), p <0.001]. Interestingly, treatment failure, death, loss to follow-up, and proportion of patients not evaluated at the end of treatment were significantly higher for the tertiary healthcare facility. Furthermore, outcome groups of TB treatment statistically significantly [FET (df) = 151.6(5), p < 0.001] differ across the primary and tertiary healthcare facilities. The cure rate (59.9% versus 40.1%) was highest in the PHC. However, rates for treatment failure (63.6% versus 36.4%), death (97.7% versus 2.3%), loss to follow-up (82.9% versus 17.1%), as well as patients not evaluated (86.2% versus 13.8%) were observed to be higher in the tertiary healthcare facility. This is contrary to the findings of a study conducted by Joseph and colleagues that found lower death and treatment failure rates in tertiary healthcare facilities [6]. Ideally, tertiary healthcare facilities are reserved for specialized care given the highly trained human resources for health who are arguably knowledgeable in managing complicated, life-threatening situations and providing pharmaceutical care that improves patients’ adherence to care [6, 28] more so that they represent major referral centres and training institutions. The findings of this research, however, cast doubt on this notion since PHC, which is purportedly a resource constraint, had a better treatment outcome. On the other hand, the PHC is located within the community, with a mechanism for default tracking, social networking, and outreach that can serve to reinforce adherence to medicines, link patients to care and modify behaviours. Additionally, as a platform for task shifting, the health workers in the PHCs possibly contribute to strict compliance with treatment guidelines which allows for more qualitative care that improves treatment outcomes for the patients. Importantly, PHC had a lower rate of failure, default, and death, further amplifying this observation. Nevertheless, lower TSR was reported in Nigeria; for instance, 34% and 37% were reported in a Nigerian teaching hospital in Kebbi State, and 17% were reported in private healthcare facilities in Lagos [29, 30, 31].

Additionally, the trend of treatment success revealed a marked decline in successful treatment outcomes over time from 68.8% in 2016 to as low as 57.6% in 2019. This drop is comparable to the findings reported in Plateau State, Nigeria [32]. Treatment success differs between primary and tertiary healthcare facilities. The tertiary healthcare facilities had the highest unsuccessful treatment rates compared to the PHC. Being HIV positive itself predicted less likelihood of successful treatment outcomes, perhaps explaining the lower successful treatment rates observed in this research. This finding concurs with a report in Lagos State, Nigeria [31].

Recent evidence indicates that knowledge of one’s HIV status is significantly associated with successful treatment outcomes for TB [30]. This is, however, inconsistent with the findings of this study, wherein only diagnostic criteria interacted statistically significantly with successful treatment outcomes. Indeed, being experienced in managing chronic diseases such as HIV is expected to instil good adherence behaviour that should be translated to TB care to help achieve successful treatment outcomes for HIV/TB coinfected patients. However, it is unclear why PLWH who were active on ART had a lower TSR than those not active on ART. This suggests that being active on ART does not translate to adherence to ART and is therefore limited in describing pill-taking behaviour.

The overall treatment success recorded in this study is higher than most of the studies conducted in the African subregion [33, 34, 35, 36]. The high treatment success recorded in this study may be attributed to the prudent implementation of the WHO treatment guidelines. Conversely, the body of literature has demonstrated adverse treatment outcomes of TB among PLWH [33, 37, 38]. The low TSR in this study could be attributed to several reasons, such as marked immunosuppression and drug interactions between anti-TB drugs and antiretrovirals [39, 40, 41]. Given the complexities of the pill burden associated with both HIV and TB treatment, adherence could be problematic.

Being treated in PHC statistically predicted successful treatment outcomes, in line with a study conducted in Ogun State, Nigeria and another similar study in South Africa that revealed that patients down-referred to PHC experienced a higher treatment success rate and lower mortality despite the likelihood of defaulting [22, 42]. However, this is contrary to the findings of a study in a resource-limited setting that identified younger age and smear positivity as independent predictors of successful treatment [25]. A study in Plateau State, Nigeria, also identified gender, age, year of enrolment and HIV status as predictors of treatment success [32].

It is a general observation that males had the highest prevalence of TB compared to females due to social habits, practices, and occupation [43]. However, this study found that more females with HIV coinfection had TB contrary to the findings of previous studies [32, 43, 44, 45]. Because this research centred on HIV/TB coinfected patients, a possible explanation for this trend is the female preponderance nature of HIV disease, which typified the findings of this study. An intervention study at the community level found higher TB prevalence among women in Ethiopia irrespective of HIV status [46].

The findings of this research revealed that in the PHC, most patients were diagnosed based on clinician judgment and acid-fast bacilli (AFB); however, chest X-ray and GeneXpert were frequently exploited for diagnosis at tertiary healthcare facilities. Perhaps the low capacity to interpret X-ray films at the PHC could account for the less frequent use of these diagnostic approaches. Comparison of treatment outcomes with diagnostic criteria revealed evidence for the relationship between successful treatments. Being diagnosed based on clinician judgment and AFB statistically interacted with successful treatment. Similarly, the cure rate was highest for patients diagnosed with AFB, clinical judgment, and even GeneXpert. This agrees with a study conducted in private healthcare facilities in Lagos, which found that being diagnosed with GeneXpert is more likely to yield an unfavourable outcome [31]. Similarly, no effect of GeneXpert when compared with sputum microscopy was observed in the mortality rate of PLWH on ART from a randomized control trial conducted in Ethiopia [47]. Furthermore, there was no significant relationship between age and successful treatment rate. This refutes the findings of Oladimeji and colleagues [31].

### Limitations of the Study

The study acknowledged certain limitations. Missing records in the healthcare facilities; poor documentation was daunting in the tertiary healthcare facilities. Among the parameters measured, treatment outcomes were the most important. Unfortunately, this measure suffered the most from missing data because of poor documentation practices. Attempting to draw insights into patients’ HIV disease stage, presence of other comorbidities, and immunological and viral load was also difficult.

Although the facilities were selected based on the high patient volume, it remains possible that patients who completed treatment elsewhere were inadvertently excluded. It was also difficult to determine whether the patients evaluated accessed both HIV and TB care in the same facilities for the study.

## Conclusion

The overall treatment success rate of TB among PLWH was low. Within the outcome categories, the proportion of patients who completed treatment and those cured compared better with unfavourable outcomes. The unsuccessful rate for TB treatment among PLWH was highest in tertiary healthcare facilities compared to PHCs. Even though being treated in the tertiary healthcare facility predicted successful treatment outcomes, treatment failure, death, and loss to follow-up were highest in the tertiary healthcare facilities.

## Recommendations

1. The government and stakeholders should strengthen healthcare facilities to support comprehensive HIV/TB care through a sustainable funding system.
2. Appropriate interventions that would strengthen the treatment supporter and health system through medication education and adherence counselling should be emphasized at tertiary healthcare facilities.
3. There is a need for a comprehensive strategy that allows holistic management that incorporates disease management and psychosocial and structural factors that may influence treatment outcomes among HIV/TB coinfected patients in order to improve treatment outcomes.

## Contribution to Knowledge

The study contributes to knowledge by providing evidence for cascading TB treatment to PHCs in Nigeria. The findings of this research will guide policy direction on the need to also strengthen PHC structures to drive the attainment of the end TB goals in Nigeria. The study has expanded the frontiers of knowledge by providing information and data on the performance of primary and tertiary healthcare facilities and outcomes of TB, specifically among PLWH for policymakers to strengthen collaborative TB/HIV care.

## Directions for Future Research

The major limitation of this study was missing data, while Markov chain Monte Carlo (MCMC) simulation, a robust statistical method of handling missing data would have been worthwhile and outcome measures of this nature would best be investigated as such without any form of statistical manipulation. It is hereby recommended that future research of this nature should combine qualitative extrapolations on a small subset of TB/PLWH to address these limitations. Similarly, adherence-related research could provide useful information on important factors militating against the achievement of a high treatment success rate.

## Data Availability

The ethical review boards of the facilities used in the research emphasise the confidentiality of the data. Hence, making the data available is difficult, unless otherwise on request.

## Declarations

## Abbreviations

AIDS: Acquired Immune Deficiency Syndrome
ART: Antiretroviral therapy
DASH, Lafia: Dalhatu Araf Specialist Hospital, Lafia
ERCC Hospital, Alushi: Evangelical Reformed Church of Christ Hospital, Alushi
FET: Fisher’s Exact Test
FMC, Keffi: Federal Medical Centre, Keffi
HIV: Human Immunodeficiency Virus
NACA: National Agency for the Control of AIDS
NAIIS: Nigeria HIV/AIDS Indicator and Impact Survey
NTBLCP: National Tuberculosis and Leprosy Control Program
PHC: Primary Healthcare Centre
PLWH: People Living with Human Immunodeficiency Virus
TB: Tuberculosis
TSR: Treatment Success Rate
UNAIDS: Joint United Nations Program on HIV/AIDS
WHO: World Health Organization

## Ethics approval and consent to participate

The conduct of this research was approved by the Health Research and Ethics Committee of the Federal Medical Centre Keffi with the reference number FMC/KF/HREC/384/20. Confidentiality was ensured and observed throughout the study. No personal identifiers were used during the data collection and patient information was treated with the utmost confidentiality.

## Consent for publication

Not Applicable.

## Availability of data and materials

The datasets used and/or analysed during the current study are available from the corresponding author on reasonable request.

## Competing interests

The authors declare that they have no competing interests.

## Funding

No funding was received for the conduct of this study.

## Notes

### Competing Interest Statement

The authors have declared no competing interest.

### Funding Statement

No funding was secured for this research work. The research was funded by the authors

### Author Declarations

1. Health Research and Ethics Committee of Federal Medical Centre Keffi, Nasarawa State 2. ERCC Health Centre, Alushi, Nassarawa Eggon Local Government Area, Nasarawa State

